# Improving older adults’ vaccination uptake: are existing measures of vaccine hesitancy valid and reliable for older people?

**DOI:** 10.1101/2021.09.29.21263161

**Authors:** Nicola Cogan, Allyson J. Gallant, Louise A. Brown Nicholls, Susan Rasmussen, David Young, Lynn Williams

**Affiliations:** School of Psychological Sciences and Health, University of Strathclyde, 40 George Street, Glasgow, G1 1QE; Faculty of Health, Dalhousie University, 5968 College Street, Halifax, Nova Scotia, Canada, B3H 4R2; Department of Mathematics and Statistics, University of Strathclyde, 26 Richmond Street, Glasgow, G1 1XH

**Keywords:** vaccine hesitancy, ageing, influenza, pneumococcal, shingles, psychometrics

## Abstract

Older adults are particularly vulnerable to vaccine-preventable diseases (VDU), due to decreased immunity and increased comorbidity. Vaccination can support healthy ageing and help reduce morbidity, mortality, and loss of quality of life associated with VPDs. Despite the availability of effective vaccines, many countries, including the UK, fail to reach recommended coverage levels. Psychosocial factors are recognised as providing important insights into the determinants of vaccination uptake. Little research has sought to establish psychometrically sound scales of vaccine attitudes with older adults. In the present study, a total of 372 UK-based participants (65-92 years, *M* = 70.5 yrs, *SD* = 4.6) completed a cross-sectional, online survey measuring health and socio-demographic characteristics in relation to vaccination uptake for influenza, pneumococcal and shingles. Two recently developed vaccination attitude scales, the 5C scale and the Vaccination Attitudes Examination (VAX) scale, were also administered to test their reliability and validity for use with an older adult population. Additional scales used to examine convergent and discriminant validity, the Beliefs about Medicines Questionnaire, the Perceived Sensitivity to Medicines Scale, the Medical Mistrust Index, the Perceived Stress Scale, and the Interpersonal Support Evaluation List, were included. The factor structure of the 5C and VAX scales was confirmed. Both scales showed good internal reliability, convergent, discriminant and concurrent validity, supporting their use with older adult populations. The 5C and VAX scales were found to be reliable and valid psychosocial measures of vaccine hesitancy and acceptance within a UK-based, older adult population. Future research could use these scales to evaluate the impact of psychological antecedents of vaccine uptake, and how concerns about vaccination may be understood and addressed among older adults.

**Ethics approval & informed consent:** Ethical approval (34/26/11/2019/Staff Williams) was granted by the School of Psychological Sciences and Health Ethics Committee, University of Strathclyde. (SEC19/20: Williams, Nicholls, Rasmussen, Young & Gallant). Approved on 8^th^ January 2020.

## Introduction

Outbreaks of vaccine-preventable diseases (VPDs) are a growing international concern particularly in the current COVID-19 pandemic (Suryadevara, 2021). As a result of these outbreaks there is an urgent need to focus on the development and evaluation of interventions to increase vaccine uptake to protect the expanding demographic of the older adult (aged ≥ 65 years) population (Privor-Dumm et al., 2020; Williams et al., 2020). Due to decreased immunity and greater likelihood of existing chronic health conditions, older adults are more susceptible to infectious diseases and have altered immune response to vaccinations (Coll et al., 2020; Russell et al., 2018). Vaccination can support healthy ageing and efforts to increase uptake among older adults seek to reduce morbidity, mortality, and loss of quality of life associated with VPDs (Ozawa et al., 2016). Vaccination uptake varies by vaccine (Klett-Tammen et al., 2015); despite the availability of effective vaccines, many countries fail to reach recommended coverage levels (Doherty et al., 2018). Within the UK, older adults (aged ≥ 65 years) are freely offered an annual influenza vaccine, a single-dose pneumococcal vaccine and a single-dose shingles (aged 70-79 years) vaccine. However, uptake has been found to fall below the World Health Organization’s (WHO’s) target of 75% (Dios-Guerra et al., 2017; Sheikh et al., 2018). For influenza vaccination, between 2004-05 and 2019-20, uptake has fluctuated between 71-75% among older adults (Health Protection Scotland, 2021b).Pneumococcal vaccination uptake is typically around 51-69% (Briggs et al., 2019; Frank et al., 2020), and shingles uptake rates can be lower than 50% (Health Protection Scotland, 2021a).

Given these uptake rates, it is important to understand the reasons for low uptake and/or vaccine hesitancy. A recent population-level cohort study of UK adults (aged ≥ 65 years) found that uptake for influenza, shingles and pneumococcal vaccination are patterned by ethnicity, deprivation, household size and comorbidities (Tan et al., 2021). In addition, psychosocial factors are increasingly being recognised as providing important insights into the determinants of vaccination behaviour (Betsch et al., 2018, 2020; Schmid et al., 2017). Measuring vaccination-related psychosocial factors is necessary to identify target populations, determine potential reasons for under-vaccination and inform the design of tailored and cost-effective interventions (Butler & MacDonald, 2015).

Recently, two self-report measures, the 5C scale (Betsch et al., 2018) and the Vaccination Attitudes Examination (VAX) Scale (Martin & Petrie, 2017), have been developed to help identify reasons why people do or do not vaccinate. The 5C scale is a novel measure of five psychological antecedents, or precursors, of vaccination behaviour: confidence, complacency, constraints, calculation, and collective responsibility. The VAX Scale assesses attitudes that may underlie vaccine hesitancy across four domains: mistrust of vaccine benefit, worries about unforeseen future effects, concerns about commercial profiteering, and preference for natural immunity. These measures are useful for predicting vaccination behaviour; however, to date, they have yet to be tested in older adults aged ≥ 65 years. While the 5C scale has been tested once with slightly older adults, this sample was only middle aged (*M* = 48 years). The VAX scale has been tested with young adults only (typically those in their 20’s and 30’s) and, therefore, it is not known if it can distinguish between vaccinating and non-vaccinating older adults.

Presently, this study aimed to examine health and socio-demographic characteristics in relation to vaccination uptake among UK-based older adults (aged ≥ 65 years) for influenza, pneumococcal and shingles. It sought to test the psychometric properties of the 5C (Betsch et al., 2018) and the VAX (Martin & Petrie, 2017) and use confirmatory factor analysis to elucidate the underlying factor structures of these measures, thereby supporting their generalisability with an older adult population. Further, it aimed to establish the psychometric properties of these measures including their internal consistency and their discriminant and convergent validity. Building upon earlier work assessing independent predictors of older adults’ vaccination uptake (Nicholls et al., 2021), the concurrent validity of the measures was also examined.

## Methods

### Participants

Participants were 372 older adults aged 65 to 92 years (*M* = 70.5 yrs, *SD* = 4.6). Participants were living in the UK, generally in good health, not diagnosed with a neurological condition, and living independently in the community (see Table 1 for socio-demographic characteristics).

**Table 1.** Participants’ socio-demographic data

| **Variables** | **n (372 total)** |
| --- | --- |
| Age | $M = 70.5$ ($SD = 4.6$) |
| Gender |  |
| Female | 184 (49.7%) |
| Male | 184 (49.7%) |
| Prefer not to say | 2 (.5%) |
| Marital Status |  |
| Married | 248 (66.8%) |
| Widowed | 47 (12.7%) |
| Separated/Divorced | 35 (9.4%) |
| Co-habiting | 21 (5.7%) |
| Single | 20 (5.4%) |
| Ethnicity |  |
| White-British | 360 (97%) |
| White-Other | 9 (2.4%) |
| Asian | 1 (.3%) |
| Mixed/Multiple | 1 (.3%) |
| Self-Rated Overall Health |  |
| Very Good | 125 (33.6%) |
| Good | 201 (54%) |
| Fair | 41 (11%) |
| Poor | 5 (1.3%) |
| Education |  |
| High School | 65 (17.5%) |
| College | 114 (30.6%) |
| University | 109 (29.3%) |
| Postgraduate | 84 (22.6%) |
| Deprivation Quintile |  |
| 1 (Most Deprived) | 57 (16.3%) |
| 2 | 83 (23.8%) |
| 3 | 69 (19.8%) |
| 4 | 61 (17.5%) |
| 5 (Least Deprived) | 79 (22.6%) |

NB: % calculations exclude missing data

### Procedure

An online, cross-sectional survey was administered using Qualtrics. Data collection took place between 8^th^ February and 17^th^ March 2020, prior to the first COVID-19 lockdown in the UK. Participants were recruited through university participation panels and social media (e.g., Twitter and Facebook).

### Measures

The online survey included the 5C scale (Betsch et al., 2018) and the VAX scale (Martin & Petrie, 2017). The Beliefs About Medicines Questionnaire (BMQ;Horne et al., 1999), the Perceived Sensitivity to Medicines scale (PSM; Horne et al., 2013), the Medical Mistrust Index (MMI; LaVeist et al., 2009), the Perceived Stress Scale (PSS; Cohen et al., 1994), and the Interpersonal Support Evaluation List (ISEL-12; Cohen et al., 1985) (see supplementary material for further scale information) were also included. Participants were asked their age, gender, marital status, ethnicity, self-reported overall health, education, and socio-economic status. Participants were also asked whether they had received the influenza vaccination in the past 12 months and if they had ever received the pneumococcal vaccination. Those aged 70 and older were asked about previous uptake of the shingles vaccination. Self-reported vaccine status has been found to be a sensitive and fairly specific indicator of actual vaccine status (Irving et al., 2009).

### Data analyses

Analyses were performed using SPSS version 23. First, internal consistency using MIIC rather than Cronbach’s α was used due to the low number of items in each subscale. Next, to verify the structural validity of the 5C and VAX scales, confirmatory factor analysis (CFA) was performed. The following indices of model fit were considered: comparative fit index (CFI), the Tucker Lewis Index (TLI), root mean square error of approximation (RMSEA), and standardised root mean square error of approximation and MIIC. CFI and TLI values between .90 and .95 and RMSEA values between .05 and .08 are indicative of acceptable model fit (Chen, 2007; Hu & Bentler, 1999; Kline, 2005). Convergent validity were examined using the correlations between the 5C and VAX with the MMI, BMQ, and PSM. Discriminant validity was assessed by examining the correlations between the 5C and VAX with the PSS and IESL-12. Finally, concurrent validity was assessed using logistic regressions (correct classification rates) within each scale, to determine if they successfully predicted vaccination behaviour (dependent variable: vaccinated yes/no for influenza, pneumococcal and shingles). All tests were two-tailed, and *p* < 0.05 was considered statistically significant.

## Results

### Internal reliability

For the 5C, good internal consistency was found across all subscales (with MIIC > 0.15): confidence, MIIC = 0.74; complacency, MIIC = 0.30; constraints, MIIC = 0.47, calculation MIIC = 0.58, and collective responsibility MIIC = 0.32. For the VAX, a total mean score MIIC = 0.45 was obtained with subscales: mistrust of vaccine benefits, MIIC =.*74;* worries over unforeseen future effects, MIIC =.*49;* concerns about commercial profiteering, MIIC =.73; and preference for natural immunity, MIIC =.67; all demonstrated good internal consistency.

### Factor structure of 5C and VAX

The factor structure of the 5C and VAX was assessed using CFA. CFA was run on all 5C subscales and the 12 VAX items grouped into four subscales (three items per subscale). For the tested models, RMSEA values were reasonable (values near to 0.08). The CFI and TLI were above the acceptable value of 0.90 (5C: CFI = 0.959, TLI = 0.946; VAX: CFI = 0.978, TLI = 0.970) suggesting good fit and verifies the structural validity of the scales when tested with an older adult population.

### Convergent and discriminant validity

Table 2 shows the correlations measuring associations between the mean 5C subscale scores, VAX total score and MMI, BMQ and PSM total means for the sample. Mean 5C subscale scores and VAX total scores correlated positively with MMI, BMQ, and PSM, showing good convergent validity. Discriminant validity was assessed by examining the correlations between the mean 5C subscale scores and VAX total score with PSS and IESL-12. As expected, these correlations were found to be weaker than those observed with the MMI, BMQ, and PSM.

**Table 2.**
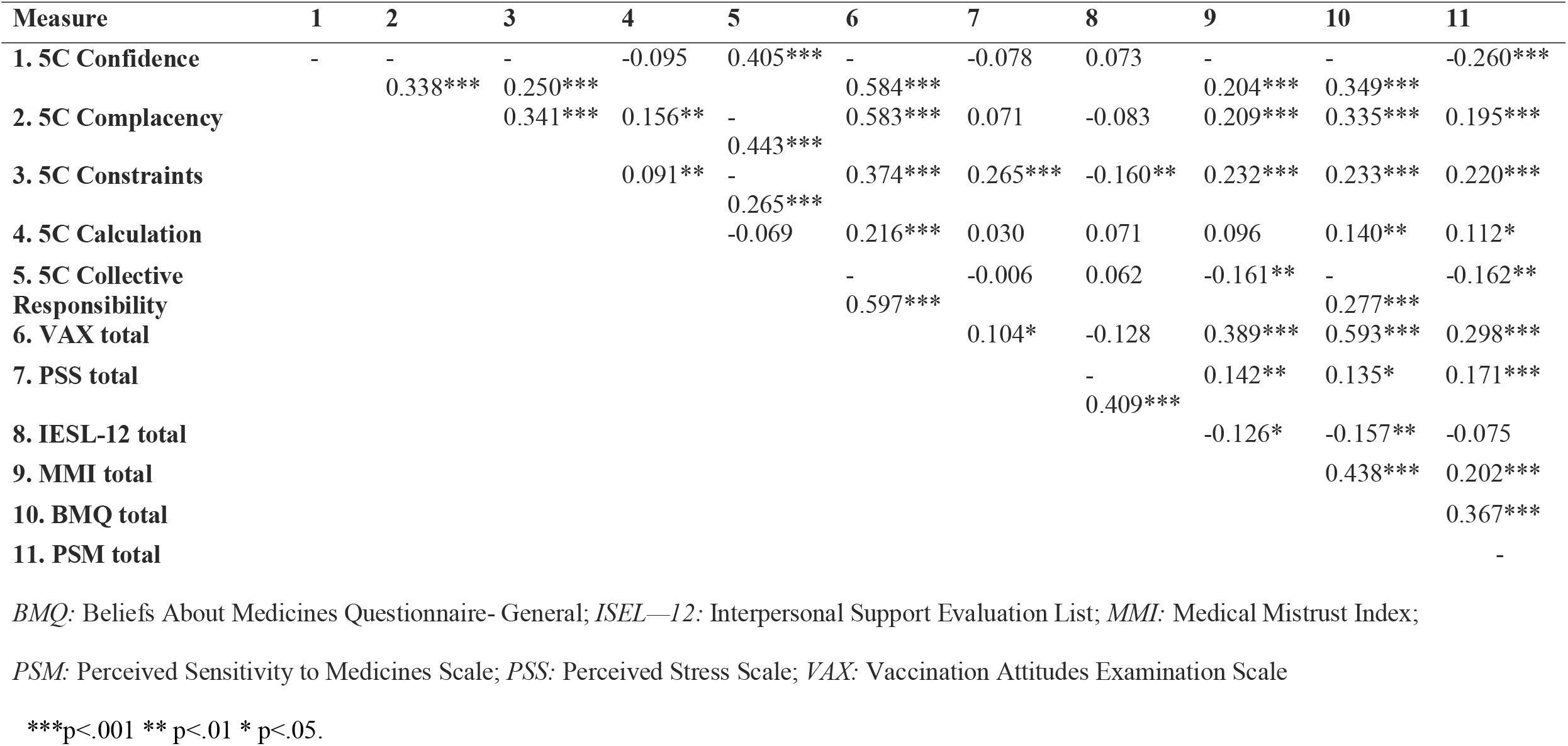
Correlation matrix of associations between mean VAX total and 5C scores and relevant questionnaires to test convergent and discriminant validity

### Concurrent validity

Concurrent validity was assessed using logistic regression to determine if the 5C and VAX scales successfully predicted vaccination behaviour (vaccinated yes/no) for influenza, pneumococcal, and shingles. Multivariate logistic models were constructed to determine the independent predictors of lack of vaccine uptake for each vaccine. Results (see Table 3) showed that both the 5C and VAX scales were able to correctly classify vaccinators and non-vaccinators across each of the vaccines, thus showing good concurrent validity.

**Table 3.**
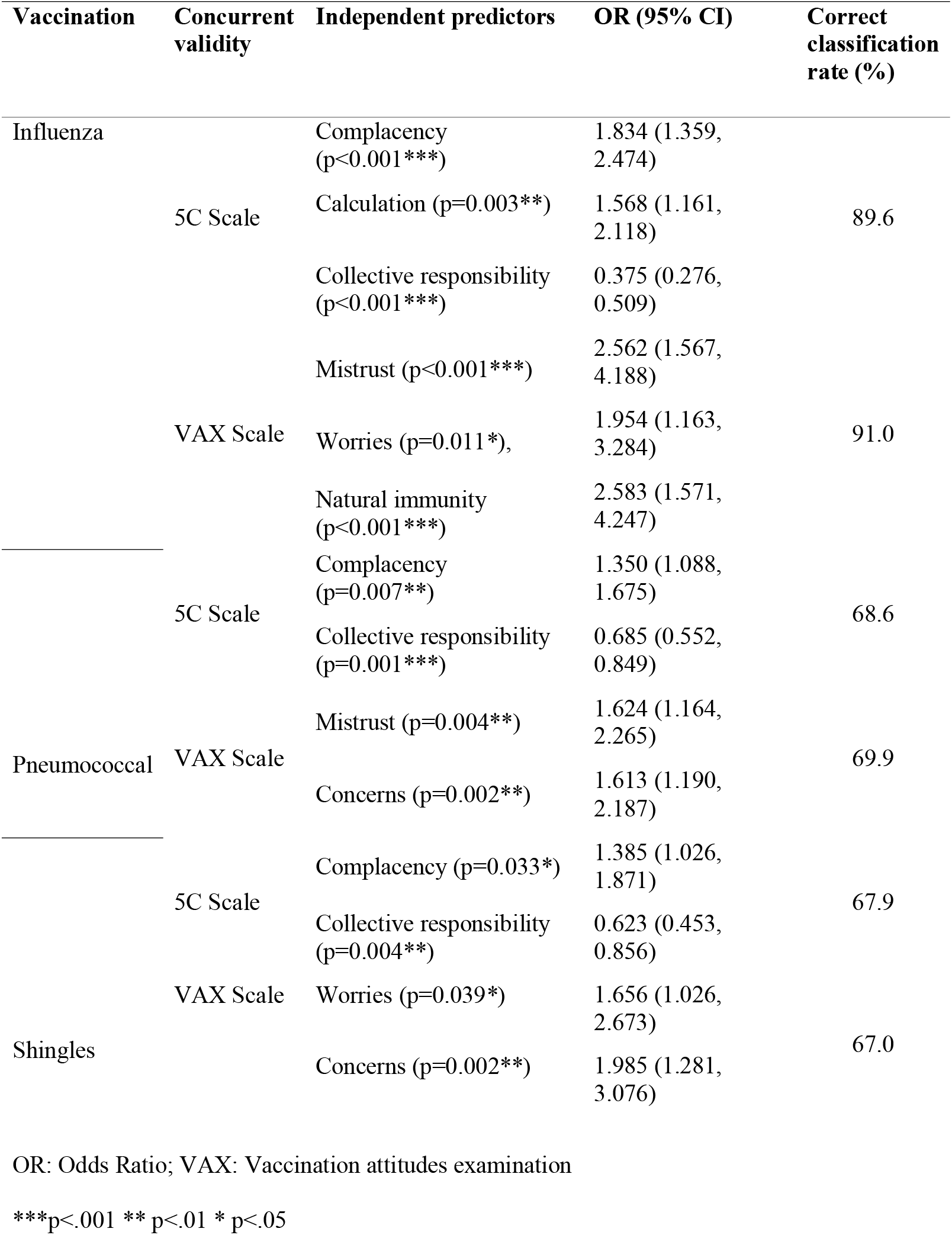
Multivariate logistic regression analyses within the 5C and VAX Scales for predicting older adults’ non-uptake of influenza, pneumococcal, and shingles vaccines.

### Discussion

This study aimed to establish the reliability and validity of the 5C (Betsch et al., 2018) and VAX (Martin & Petrie, 2017) scales, two psychosocial measures of vaccine uptake, within a UK older adult population. Results revealed satisfactory psychometric properties for use with older adults as the results were indicative of statistically sound factor models. The 5C and VAX were shown to have good internal consistency and good levels of convergent and discriminant validity. As reported in earlier work, psychosocial factors independently predicted older adults’ hesitancy towards the influenza, pneumococcal, and shingles vaccines (Nicholls et al., 2021). Presently, these findings show that the two scales have good concurrent validity.

The findings contribute to metrics which aim to assess vaccine hesitancy (Betsch et al., 2018; Huza, 2020; Martin & Petrie, 2017; Wood et al., 2019) through demonstrating that both the 5C and VAX are reliable and valid measures for use with an older adult (65+) population. Such measures can be used to inform targeted public health action to increase vaccine uptake with older adults, using appropriate strategies, policies, and interventions to reduce vaccine hesitancy (Frank et al., 2020; Jarrett et al., 2015; Nicholls et al., 2021). Given the pressures on healthcare systems during the COVID-19 pandemic, having sound measures of vaccine hesitancy with older adults, who may be more vulnerable to adverse outcomes, has come into strong focus (Roller-Wirnsberger et al., 2021).

Future studies are recommended to determine the relationship between psychosocial and other factors associated with older adults’ vaccine uptake using the 5C and VAX scales. Given that participants in the current study were high functioning and were living independently at home with minimal assistance, the findings may not be generalisable to older adults with lower functional abilities, for example, those who are experiencing cognitive impairment or limitations in everyday functioning (Nicholls et al., 2021). Future work could therefore explore the psychometric properties and feasibility of the 5C and VAX in more diverse populations of older adults (e.g., ethnic minorities, varied functional levels) and socio-cultural contexts (e.g., low-income countries). Longitudinal studies are also required to establish their ability to predict outcomes such as objective measurements of vaccination uptake relative to the total number of recommended dosages. Furthermore, it is possible that participants may report less vaccine hesitancy face-to-face than through an anonymous online survey, which may encourage self-disclosure on sensitive items (Hollier et al., 2017). It would be useful for future work to evaluate possible differences in vaccine hesitancy using the 5C and VAX using online surveys, telephone surveys, and face-to-face assessments. Given that older adults are at increased risk of disease morbidity, including VPDs, targeting research to inform intervention programmes to increase uptake, using psychometrically validated measures of vaccine hesitancy, is of paramount importance. This is particularly pertinent given the global challenges associated with the dynamic and changing nature of COVID-19, highlighting the need for ongoing research on vaccine hesitancy (Holeva et al, 2021).

In conclusion, the 5C and VAX scales were found to be reliable and valid within a UK-based, older adult (65+) population. Future research could use these scales to evaluate the impact of psychological antecedents of vaccine uptake, and how concerns about vaccination may be challenged and reversed in older adults. Improved measurement and targeted education around VPDs, disease risk, and vaccine benefits may be required to increase vaccine coverage. It is hoped that these findings will influence future research and intervention development with older adults aimed at achieving this. The 5C and VAX scales appear to be useful measures to help understand the health and vaccination attitudes that promote and deter older adults’ vaccination uptake.

## Supporting information

Supplementary file of measures used

Evidence of ethical approval granted

## Data Availability

The data can be made available upon request

## Acknowledgements

This work was supported by the Chief Scientist Office, Scottish Government [grant number CGA/19/52].

## Declarations

### Statement Regarding Informed Consent

Informed consent was obtained from all individual participants included in the study.

### Statement Regarding Ethical Approval

Ethical approval (34/26/11/2019/Staff Williams) was granted by the School of Psychological Sciences and Health Ethics Committee, University of Strathclyde. (SEC19/20: Williams, Nicholls, Rasmussen, Young & Gallant). Approved on 8^th^ January 2020. All procedures performed in studies involving human participants were in accordance with the ethical standards of the institutional and/or national research committee and with the 1964 Helsinki declaration and its later amendments or comparable ethical standards.

## Notes

**Conflicts of interest declaration:** The authors declare that they have no conflict of interest.

### Competing Interest Statement

The authors have declared no competing interest.

### Author Declarations

Ethical approval (34/26/11/2019/Staff Williams) was granted by the School of Psychological Sciences and Health Ethics Committee, University of Strathclyde. (SEC19/20: Williams, Nicholls, Rasmussen, Young & Gallant). Approved on 8th January 2020"

