## Supplementary file of measures used for "Improving older adults’ vaccination uptake: are existing measures of vaccine hesitancy valid and reliable for older people?"

**Supplementary materials**

### **Further scale information**

For the convergent validity, we included the following scales: Beliefs about Medicines Questionnaire (BMQ) by Horne et al., (1999) the Perceived Sensitivity to Medicines Scale (PSM) by Horne et al., (2013) and the Medical Mistrust Index (MMI) by LaVeist et al., (2009).

The 8-item general version of the BMQ was included and asks participants about their personal views, beliefs and worries about taking medicines (e.g., “Doctors use too many medicines”). Responses are indicated on a 5-point Likert answer option, which varies from 1 ‘strongly disagree’ to 5 ‘strongly agree’. Higher scores indicate stronger and more negative beliefs about the use of medicines (10). This scale was selected to probe perceptions about medicines in general and evaluate similarities to vaccination attitudes.

The short 5-item PSM scale was included to understand in what way participants generally respond when they take prescribed medication (e.g., “My body is very sensitive to medicines”). Responses are captured on a 5-item Likert-type scale and range from 1 ‘strongly disagree’ to 5 ‘strongly agree’. Higher scores indicate more perceived sensitivity to medicines and someone with a high score may less willing to take as much medication to either avoid a potential adverse effect or because they believe they require less medication compared to others with lower PSM scores. Here, we wanted to see if these perceptions about medicines were similar to attitudes about vaccination.

The 7-item MMI asks participants about their feelings and trust with regards to the National Health Service (the publicly funded national healthcare system in the U.K.) in their local area including a hospital, clinic or the general health care system (e.g., “mistakes are common in the NHS”). Responses are captured on a 5-item Likert-type scale and range from 1 ‘strongly disagree’ to 5 ‘strongly agree’. Higher scores indicate more perceived mistrust in the health service and these negative believes may be similar in those with anti-vaccination attitudes.

For discriminant validity we included the following scales: Perceived Stress Scale (PSS; Cohen et al., 1994) and the Interpersonal Support Evaluation List (ISEL-12; Cohen et al., 1985).

A 4 item version of the PSS assessed potentially stressful events over the previous month. Items in the scale are scored on five point scale (0=never, 4= very often) and help identify how different events can affect emotions and perceptions of stress (e.g. In the last month, how often have you felt that you were unable to control the important things in your life?). Higher scores from this scale indicate higher perceived stress levels.

The 12 items of the ISEL-12 assessed social support across the subscales of availability of appraisal (e.g. ‘there is someone I can turn to for advice about handling problems with my family’), belonging (e.g. ‘if I decide one afternoon that I would like to go to a movie that evening, I could easily find someone to go with me’), and tangible help/assistance (e.g. ‘if I were sick, I could easily find someone to help me with my daily chores’). Responses are captured on a 4-point scale (1 = ‘definitely false’ to 4 = ‘definitely true’) and totalled to give a maximum of 12 for each subscale, and 36 for overall social support, with higher scores indicating more perceived support.

### **Confirmatory factor analysis**

Confirmatory factor analysis of the four previously determined VAX subscale factors was achieved using the Lavaan package for R version 3.4.3. A diagonally weighted least squares algorithm specifying ordinal data was employed using all 12 VAX items in the following model and fit*:

model: <- ‘

subscale1 =~ vax1r + vax2r + vax3r

subscale2 =~ vax4 + vax5 + vax6

subscale3 =~ vax7 + vax8 + vax9

subscale4 =~ vax10 + vax11 + vax12

fit <- cfa(model, data = vaxdata,

ordered=c("vax1r","vax2r","vax3r",

"vax4", "vax5", "vax6",

"vax7", "vax8", "vax9",

"vax10", "vax11", "vax12"))

* Note: subscale1 = trust of benefits, subscale2 = worries over future effects, subscale3 = concerns about commercial profiteering and subscale4 = preference for natural immunity, reverse item scoring is indicated by (r).

Results of confirmatory factor analysis suggest a good model fit with *X*^2^ (48, *N* = 243) = 33.898, *p* = 0.938, Comparative Fit Index = 1.00, Normed Fit Index = 0.998, Tucker-Lewis Index = 1.001, Root Mean Square Error of Approximation 0.000, and 90% confidence interval = 0.000 – 0.010.
