## Supplementary material for "Improving older adults’ vaccination uptake: are existing measures of vaccine hesitancy valid and reliable for older people?": Evidence of ethical approval granted

8th January 2020

Dear Applicants,

I can confirm that the School of Psychological Sciences and Health Ethics Committee (SEC) has approved this study.

The SEC must be informed of any changes you plan to make to the research project, including any staff changes should these occur.

In the event of any ethical issues that arise during the project or if any adverse events occur, please inform SEC of these events at the earliest opportunity.

Oh behalf of the Committee, I wish you success with this project.

Kind regards,

Dr Joanne Cummings
Psychological Sciences and Health Ethics Committee
University of Strathclyde
Graham Hills Building, 40 George Street,
Glasgow, G1 1QE


**The place of useful learning**
The University of Strathclyde is a charitable body, registered in Scotland, number SC015263
